# Explainable AI and public reactions to AI-involved adverse diagnostic events: a vignette study

**DOI:** 10.64898/2026.05.26.26353870

**Authors:** Jungmin Choi, Yeun Joon Kim, Pengzhao Lyu, Yingyue Luna Luan, Soo Min Toh

## Abstract

Artificial intelligence (AI) is increasingly incorporated into diagnostic decision-making, raising questions about physician responsibility following AI-involved adverse diagnostic events. Explainable AI (XAI) has been proposed to improve transparency and trust, but its influence on public reactions remains unclear.

In a randomised vignette-based experiment, 652 adults from the United States and United Kingdom were assigned to one of six conditions in a 3 (diagnostic source: AI alone, human radiologist alone, or human–AI collaboration) × 2 (explanation: present or absent) between-subjects design. Participants read a scenario in which a chest X-ray was initially interpreted as normal but lung cancer was diagnosed five months later, indicating that the original interpretation had missed the cancer. In explanation conditions, participants received additional information about how the diagnosis had been reached, including AI heatmap-based explanations in the AI conditions. Participants rated radiologist responsibility, likelihood of complaint, and intention to pursue legal action.

Among 652 participants (mean age 42.2 years; 50.2% female), responsibility ratings were significantly lower when AI alone made the diagnostic decision (mean 4.73, 95% CI 4.53–4.93) compared with human-only decision-making (5.78, 95% CI 5.59–5.98; p<0.001) and human–AI collaboration (5.54, 95% CI 5.34–5.74; p<0.001). Complaint likelihood showed a similar pattern. Intentions to pursue legal action followed the same directional trend but were marginally significant. Neither explanations nor explanation-by-source interactions were associated with outcome measures.

These findings suggest that the public expects physicians to remain accountable when AI is involved in diagnostic decision-making, particularly in collaborative settings. Providing explanatory information about how AI systems reach decisions may be insufficient to change perceptions of physician responsibility following adverse diagnostic events.

## INTRODUCTION

Physicians are often regarded by the public as the primary bearers of responsibility when adverse diagnostic events occur.^1,2^ The integration of artificial intelligence (AI) into clinical decision-making complicates this responsibility, as AI systems are frequently described as “black boxes” whose reasoning processes are opaque to both patients and clinicians.^3^

Explainable AI (XAI) has been proposed as a way to make AI-supported clinical decisions more transparent and trustworthy by enabling physicians to understand and scrutinize algorithmic outputs and communicate them to patients.^4^ Although initially developed for clinician oversight, XAI is increasingly recognized for its potential importance for fostering transparency and trust among patients.^5^ However, it remains unclear whether XAI influences public reactions following AI-involved adverse diagnostic events. This study therefore investigates whether XAI alters reactions toward physicians following adverse diagnostic events involving AI.

## METHODS

The study was conducted with US and UK adults via Prolific. Participants were randomly assigned in one of the conditions in a 3 (Diagnostic Source: AI vs. Human vs. Human–AI Collaboration) × 2 (Explanation: Present vs. Absent) between-subjects design.

Participants read a vignette in which they experienced breathing difficulties and underwent a chest X-ray, with a radiologist explaining the procedure. Diagnostic source was manipulated such that the X-ray was interpreted by an AI system, a radiologist, or through collaboration between the two. Explanation was manipulated by providing additional information regarding how the diagnosis had been reached. In the AI–Explanation condition, participants received explanatory information describing how the AI analyzed the X-ray and visualized its decision-making process using a heat map.^6^ In the Human–Explanation condition, participants received explanatory information describing how the radiologist inspected the X-ray. In the Collaboration–Explanation condition, both forms of explanation were provided. In the No-Explanation conditions, no additional information was given.

Across conditions, the image was initially interpreted as normal, and no further treatment was recommended. At the end of the vignette, participants learned that symptoms worsened and lung cancer was detected 5 months later, indicating that the initial interpretation had missed the cancer.

Participants rated radiologist responsibility, likelihood of complaint, and intention to pursue legal action (7-point scales). Data were analyzed using 2-way ANOVA with pairwise comparisons (2-sided *p* < .05). Participants who failed manipulation checks were controlled. Written informed consent was obtained from all participants. Vignettes are available in the supplemental material. Ethics Review Group of the University of Cambridge gave ethical approval for this work (approval no. 24-28).

## RESULTS

Participants included 652 adults (327 [50.2%] female, 320 [49.1%] male, 5 [0.8%] nonbinary/prefer not to say; mean [SD] age, 42.15 [13.91] years). Responsibility ratings were significantly lower in the AI condition (4.73 [95% CI, 4.53–4.93]) compared with the Human condition (5.78; 95% CI [5.59–5.98]; *p* < .001) and the Human–AI Collaboration condition (5.54; 95% CI [5.34–5.74]; *p* < .001). The difference between Human and Collaboration conditions was marginal (*p* = .094; Table 1, Figure 1).

**Table 1.** Effects of the conditions on the outcomes.

|  | **F** | **P value** | **$\eta_p^2$** |
| --- | --- | --- | --- |
| **Responsibility** |  |  |  |
| Source of Error (0 = Human; 1 = AI; 2 = Collaboration) | 30.17 | < .001 | .09 |
| Explanation (0 = Absent; 1 = Present) | .93 | .335 | .001 |
| Source of Error × Explanation | 1.19 | .304 | .004 |
| **Complaint Likelihood** |  |  |  |
| Source of Error | 8.56 | < .001 | .03 |
| Explanation | .72 | .398 | .001 |
| Source of Error × Explanation | .65 | .523 | .002 |
| **Legal Action Intention** |  |  |  |
| Source of Error | 2.08 | .125 | .01 |
| Explanation | .25 | .616 | .00 |
| Source of Error × Explanation | .92 | .399 | .003 |

**Figure 1.**
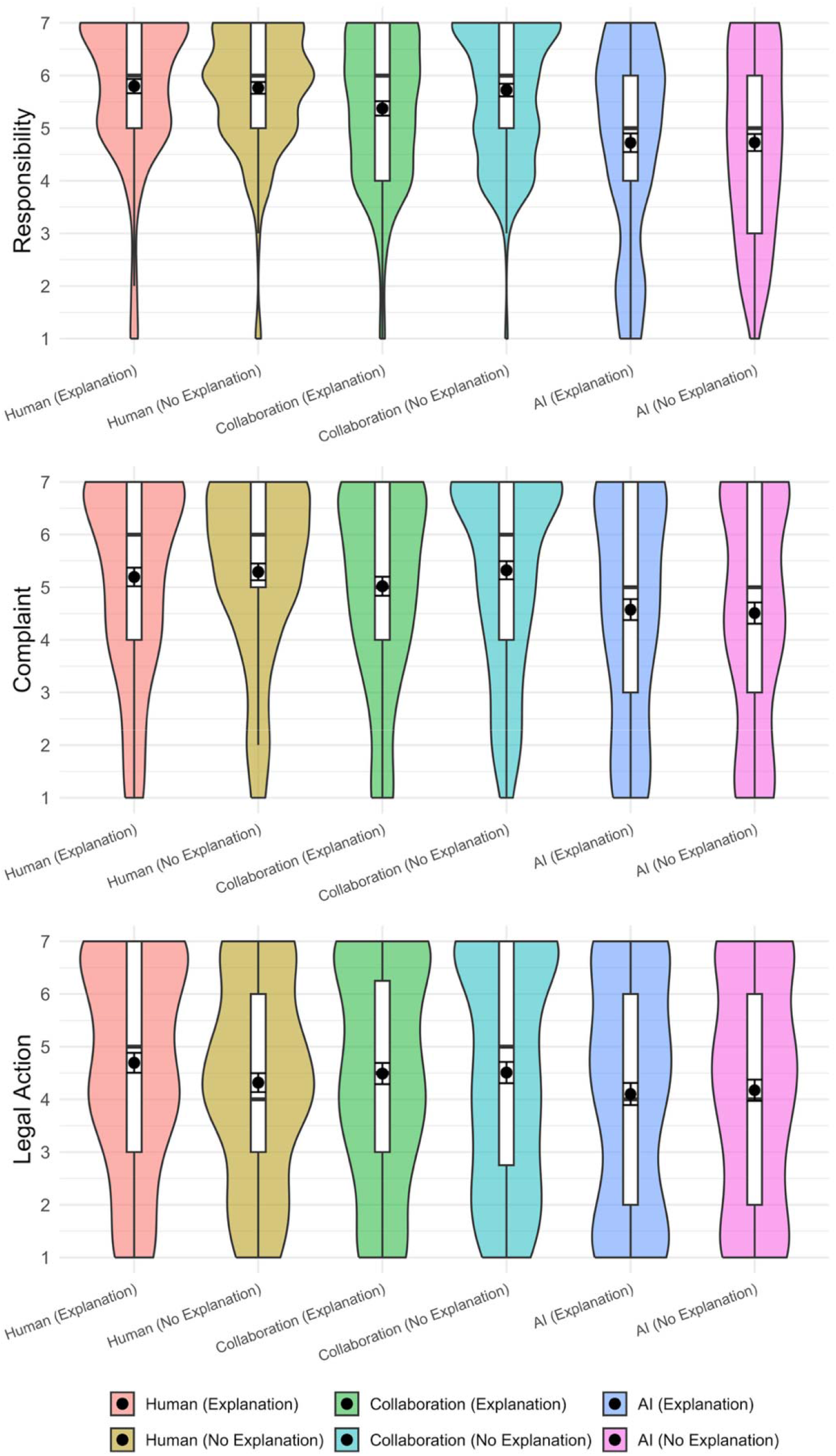
Mean ratings across conditions. Ratings are from 1 to 7. Error bars indicate standard errors of each mean rating.

Complaint likelihood followed a similar pattern: ratings were significantly lower in the AI condition (4.55; 95% CI [4.30–4.81]) than the Human condition (5.25; 95% CI [5.00–5.50]; *p* < .001) and the Collaboration condition (5.15; 95% CI [4.90–5.41]; *p* = .001), with no difference between Human and Collaboration (*p* = .595).

For legal action intention, the same pattern was observed, though the effects were marginal (AI: 4.15; 95% CI [3.87–4.42]; Human: 4.51; 95% CI [4.24–4.79]; Collaboration: 4.48; 95% CI [4.20–4.76]). Pairwise comparisons showed: AI vs Human (*p* = .065), AI vs Collaboration (*p* = .095), and Human vs Collaboration (*p* = .876).

Neither Explanation nor the Source × Explanation interaction had any effect on outcomes.

## DISCUSSION

Our findings show that the public assigns similar levels of responsibility to physicians following adverse diagnostic events occurring in human–AI collaboration and in human-only decision-making, suggesting that AI involvement does not substantially reduce expectations of physician accountability. By contrast, responsibility attributions were lower when AI was the sole decision-maker. Explanatory information—including XAI-based visual explanations—did not alter reactions, suggesting that transparency alone may be insufficient to address public concerns regarding accountability in AI-supported healthcare.^7^

Limitations include the vignette-based design and reliance on hypothetical outcomes, which may decrease the external validity of the findings. In addition, this study evaluated one visual format of XAI (heatmaps); future research may examine alternative explanatory approaches, including natural language explanations.

## CONCLUSION

Public expectations of physician accountability were similar in human-only and human–AI collaborative diagnostic decision-making. XAI did not alter public reactions following AI-involved adverse diagnostic events, indicating that explanatory information alone may have limited influence on perceptions of physician responsibility in AI-involved care.

## Data Availability

All data produced in the present study are available upon reasonable request to the authors.

## Notes

### Competing Interest Statement

The authors have declared no competing interest.

### Author Declarations

Ethics Review Group of the University of Cambridge gave ethical approval for this work (approval no. 24-28).

### Funding

- Wellcome Trust

## REFERENCES

1. Gallagher TH, Waterman AD, Ebers AG, Fraser VJ, Levinson W. Patients’ and physicians’ attitudes regarding the disclosure of medical errors. JAMA. 2003;289(8):1001–1007.

2. Blendon RJ, DesRoches CM, Brodie M, Benson JM, Rosen AB, Schneider E, Altman DE, Zapert K, Herrmann MJ, Steffenson AE. Views of practicing physicians and the public on medical errors. N Engl J Med. 2002;347(24):1933–1940.

3. Durán JM, Jongsma KR. Who is afraid of black box algorithms? On the epistemological and ethical basis of trust in medical AI. J Med Ethics. 2021;47(5):329–335.

4. Gordon L, Grantcharov T, Rudzicz F. Explainable artificial intelligence for safe intraoperative decision support. JAMA Surg. 2019;154(11):1064–1065.

5. Amann J, Blasimme A, Vayena E, Frey D, Madai VI. Explainability for artificial intelligence in healthcare: a multidisciplinary perspective. BMC Med Inform Decis Mak. 2020;20:310.

6. Rajpurkar P, Irvin J, Zhu K, et al. CheXNet: Radiologist-level pneumonia detection on chest X-rays with deep learning. PsyArXiv. https://arxiv.org/abs/1711.05225. December 25, 2017.

7. Waring JJ. Beyond blame: cultural barriers to medical incident reporting. Soc Sci Med. 2005;60(9):1927–1935.

